# What does ‘preconception health’ mean to people? A public consultation on awareness and use of language

**DOI:** 10.1101/2024.07.19.24310268

**Authors:** Danielle Schoenaker, Olatundun Gafari, Elizabeth Taylor, Jennifer Hall, Caroline Barker, Barney Jones, Nisreen A Alwan, Daniella Watson, Chandni Maria Jacob, Mary Barker, Keith M Godfrey, Emily Reason, Finlay Forder, Judith Stephenson, the UK Preconception Partnership

## Abstract

**Introduction:** There is growing scientific and policy recognition that optimising health before a potential pregnancy (preconception health) improves reproductive outcomes and the lifelong health of future children. However, public awareness on this topic is low. We conducted a public consultation to develop language recommendations and identify and prioritise approaches to inform research and improve public awareness of preconception health.

**Methods:** A public consultation was undertaken with people of any gender aged 18-50 years living in the UK who were not currently expecting a child. Public contributors were recruited through patient and public involvement, community and support groups, an existing cohort study, and an LGBTQ+ charity. An initial round of online group discussions (February/March 2021) explored public contributors’ knowledge of preconception health, their recommendations for appropriate language, and ideas about public health approaches. In a subsequent discussion round (May 2021), language recommendations were refined, and suggested approaches prioritised. Discussions were summarised based on notes taken by two researchers.

**Results:** 54 people joined the initial discussion round (66% women, 21% men, 13% non-binary or transgender; 55% aged 18-30 years, 30% 31-40 years, 15% 41-50 years). Of these, 36 people (67%) participated in the subsequent round. Very few had heard the term ‘preconception health’, understood what it means, or why and for whom it is important. They recommended avoiding unfamiliar terms without further explanation (e.g. preconception health, medical terms), using language that is positive, encouraging and gender-sensitive where possible, and using messages that are specific, non-judgmental and realistic. The phrases ‘health and wellbeing during the childbearing years’, ‘health and wellbeing before pregnancy and parenthood’ and ‘planning for parenthood’ resonated with most public contributors. School-based education, social media campaigns and the National Health Service emerged as priority approaches/settings for raising awareness.

**Conclusion:** This public consultation produced recommendations from a diverse group of people of reproductive age in the UK to improve language and prioritise approaches that increase public understanding of preconception health in ways that are relevant and appropriate to them. This should begin in schools and will require adaptation of curricula, alongside co-development of public awareness campaigns and guidance for healthcare professionals.

**Patient or public contribution:** This public consultation included a diverse group of members of the public. They were not involved in the original design of the project, but following the initial round of online group discussions, they contributed to interpretation and refinement of the emerging concepts in a subsequent round of group meetings. After the consultation activity, public contributors formed a Public Advisory Group and have subsequently been involved in other studies on the same topic. Two public contributors (ER, FF) provided critical input in the preparation and revision of this manuscript and are co-authors on the paper.

## Introduction

Preconception health describes the medical, behavioural and social risk factors of people of reproductive age (15-49 years) before or between pregnancies.^1^ These factors, such as living with diabetes or obesity, dietary habits and living circumstances, influence people’s lifelong health and wellbeing, as well as their chance of a healthy pregnancy and baby.^2^ For example, maternal folic acid supplement use reduces the risk of neural tube defects and anomaly-related terminations,^3^ while maternal and paternal obesity are associated with increased risk of obesity and the associated long-term adverse health consequences in their children.^4, 5^ Preconception risk factors are common among women and men in the UK across their reproductive years,^6^ including among those who are actively planning pregnancy,^7, 8^ and who become pregnant.^9^ Large social and economic inequalities exist in pregnancy planning and preconception health,^9–11^ and it is therefore important to apply structural changes to the systems with which people interact and provide appropriate support to enable people to be healthy.

The importance of optimising preconception health is recognised in numerous policy and public health strategies and clinical healthcare guidelines in the UK,^12^ and internationally.^13^ These strategies and guidelines suggest preconception health be addressed from individual (targeted at people planning pregnancy) and public health (reaching people across the life course irrespective of pregnancy intentions) perspectives.^2^ The UK does not currently have a national preconception care strategy or co-ordinated approach to optimising pregnancy planning and preconception health, but some local programmes are being developed, implemented and tested.^14–17^

Lack of knowledge and awareness in the general population has been a commonly reported barrier to accessing services and resources and optimise preconception health.^18–23^ In response to this, an important component of local preconception health programmes has been to provide information and raise awareness, largely focussed on information relevant to cisgender women planning pregnancy. Having access to preconception health information and being aware of steps to take to plan and prepare for pregnancy may not on its own be sufficient, but can be an important prompt for behaviour change and a key part of preconception health interventions.

To guide the development of local and national approaches to improve knowledge and raise awareness of preconception health, it is important to work with the target population through co-development of research to ensure the language and intervention platforms are relevant and appropriate. The target population – including people across their reproductive years who may become pregnant or whose reproductive partner may become pregnant – is very broad and diverse in terms of, for example, age, gender, health literacy, cultural beliefs, and intention to become pregnant or have a family. This may challenge the recruitment of study participants, the co-development and implementation of preconception health messages and interventions, and dissemination of research findings in a way that resonates with the overall target population. There is currently no published literature or guidance on language and messaging that has been co-developed with, or found to be appropriate by, the public. Moreover, while many platforms to educate and raise awareness of preconception health have been proposed by researchers, health professionals, and study participants who identify as women,^24–26^ there is a gap in perspectives from gender-diverse individuals, and the public’s preferences have not informed the basis of approaches to inform tailored co-development research on interventions and resources.

We conducted a public consultation to explore the use of language about preconception health and to inform appropriate language that is meaningful to members of the public, with a view to supporting effective preconception health research, policy and practice.

## Methods

Between February and May 2021, online group discussion meetings were conducted with people of any gender aged 18-50 years living in the UK. People were eligible to take part if they were not currently expecting a child, irrespective of whether they had children or were planning to have (more) children. To involve a diverse group of individuals, public contributors were recruited through a range of channels including: mailing lists for the University of Southampton/University Hospital Southampton’s Patient and Public Involvement (PPI) group, Health and Wellbeing Community Engagement Hub and student societies, King’s College London mental health PPI group, personal networks of the project team, Facebook local community and UK-wide support groups (e.g. for people living with diabetes, pregnancy loss or fertility treatment experience), the Pregnancy Planning Preparation and Prevention (P3) study (targeted emails to participants from ethnic minority backgrounds), and Beyond Reflections (charity supporting transgender, non-binary and questioning people, formerly ‘Chrysalis’). Anyone interested in taking part was asked to complete a Microsoft Form asking for their name, contact details, age group, gender, current pregnancy status if relevant, and availability for online group meetings.

The consultation involved two rounds (sessions) of one-hour group meetings on Zoom facilitated by the lead researcher (DS) using a topic guide, with notes taken during the discussions by a second researcher (OG or ET). One-to-one meetings were offered if this was preferred by public contributors for reasons such as availability or sensitivity of talking about the topic in a group setting. The discussions were not recorded as this was not a research project. The initial session (‘Session 1’, February/March 2021) explored public contributors’ existing knowledge about preconception health, their recommendations for appropriate language when communicating about the topic, and ideas on future public health approaches to raise awareness. Questions discussed included:

- How would you describe the time before pregnancy and why health might be important during this time?
- Have you heard of and/or what are your thoughts on the term ‘preconception health’?
- What are important things to consider when communicating about preconception health?
- What can we do to raise awareness of preconception health?

Discussions were summarised based on notes taken by the facilitator (DS) and researcher (OG). Concepts emerging from Session 1 were discussed in a meeting with the project team, including draft recommendations on language to use when communicating about preconception health (e.g. in research, the media, healthcare), and a list of suggested ways to raise awareness on the topic.

Based on this meeting, it was decided to conduct a subsequent session (‘Session 2’) of online group meetings to refine and agree on the language recommendations, apply the recommendations to an existing poster on ‘Thinking of having a baby?’ available on the Contraception Choices website,^27^ and prioritise the suggested approaches to raise awareness of preconception health. Public contributors who agreed to be contacted again were invited to Session 2 of group meetings (May 2021).

Questions discussed included:

- What are your thoughts on the recommendations for language on health and wellbeing before pregnancy and parenthood? (list of recommendations developed based on Session 1 were verbally discussed and shared on screen)
- What do you think about the language used on the poster ‘Thinking of having a baby?’ and how can it be improved based on the language recommendations we talked about?
- What do you think are the most important ways through which we should raise awareness of health and wellbeing before pregnancy and parenthood? (list of approaches identified in Session 1 were verbally discussed and shared on screen)

Discussions were synthesised and recommendations refined and updated based on notes taken by the facilitator (DS) and researcher (ET).

An anonymous poll at the end of each meeting in Session 1, and an anonymous Microsoft Form sent after each meeting in Session 1, were used to obtain feedback on public contributors’ experience of taking part. No feedback was obtained after Session 2.

## Results

Public contributors’ characteristics are outlined in **Table 1**. The initial session (Session 1) included 16 online discussion meetings (12 group meetings and 4 one-to-one meetings) with a total of 54 public contributors. The group was diverse in terms of gender identity (66% women, 21% men, 13% non-binary or transgender) and age (55% were aged 18-30 years; 30% 31-40 years; and 15% 41-50 years). All public contributors were invited to take part in the subsequent session (Session 2) which was attended by 36 people across five group meetings. The distribution of public contributors’ characteristics was similar across sessions (**Table 1**). Based on discussions during the meetings, at least one third of public contributors in both sessions were from an ethnic minority background, one third reflected on being a parent or mentioned they had children, and meetings included people with experience of physical and mental health conditions, previous pregnancy loss and complications, and varying personal intentions around future pregnancy and parenthood.

**Table 1.** Public contributor characteristics.

|  | **Session 1**<br>N = 54 | **Session 2**<br>N = 36 |
| --- | --- | --- |
| Gender identity |  |  |
| Man | 21% | 19% |
| Woman | 66% | 67% |
| Non-binary or transgender | 13% | 14% |
| Age (years) |  |  |
| 18-30 | 55% | 45% |
| 31-40 | 30% | 35% |
| 41-50 | 15% | 20% |

### Knowledge and understanding of preconception health

In Session 1, hardly any public contributors knew how to describe the time before pregnancy, how this time period could be defined, when it might start, and why or to whom it may be relevant and important. Most people did not know that health before pregnancy was important to think about. After the facilitator briefly explained that ‘women and their partners may make changes to their health and behaviours before they try to get pregnant to improve their chance of a healthy pregnancy and baby’, a follow-up question was asked to explore public contributors’ knowledge on what changes might be important to consider before pregnancy. Common thoughts shared were statements along the lines of:

> *“Trying to become pregnant is not something that is talked about, unless you are having issues getting pregnant then you go look for information”*
>
> *“I have never seen any advice on what to do and what not to do when trying to get pregnant”*

A few public contributors mentioned the importance of the mother’s age, folic acid supplement use, a healthy lifestyle including weight management, diet, alcohol and smoking, physical health conditions such as diabetes and endometriosis, and mental health and wellbeing. During the discussions, it became clear that most public contributors had some understanding that these factors may affect the ability to conceive, but no awareness of the link with health in pregnancy and beyond for parents and children. People were also surprised to hear that the health of the male partner may impact pregnancy outcomes. Public contributors who identified as men in particular thought that a healthy relationship between the future parents, and financial and housing security, were important and part of their contribution to having a family.

Few had heard the term ‘preconception health’, and very few understood what it meant or thought it may be relevant to them. The main issues they had with the term were:

- It incorrectly assumes that you know when conception is going to happen, and that it is going to happen;
- It is not specific enough and without education or further context it is unclear when, why and to whom it is relevant, or what it involves;
- ‘(Pre)conception’ is not understood by everyone, it sounds clinical, and can also have a completely different meaning (i.e. preconceived idea or bias);
- It is interpreted as referring to women’s health only and is not inclusive.

Alternative terminology was discussed; ideas included health during the fertile years, reproductive health, trying to conceive, trying to become pregnant, ready for pregnancy, thinking of having a baby, and planning to have children – but public contributors felt each alternative option had its own issues and were not entirely clear or inclusive. The phrases ‘health and wellbeing during the childbearing years’ and ‘health and wellbeing before pregnancy and parenthood’ resonated with most public contributors, and it was felt that these covered the right time period, sounded appropriate and relevant to all genders, and were easy to understand.

### Language and public health message recommendations

Important things to consider when communicating and educating people about preconception health were also discussed in Session 1 and summarised into recommendations for language and public health messages by the facilitator, which were then refined and agreed on in Session 2 (Box 1).

#### Box 1. Recommendations for language and public health messaging about preconception health.

##### Language recommendations

- Avoid the term ‘preconception health’ without further explanation or context.
- Avoid clinical and medical terms, such as conception and spina bifida, or use these terms with a simple description.
- Use positive, optimistic and encouraging language (avoid a focus on problems and what can go wrong, or explain how to reduce any risks).
- Replace strong and definitive language with more nuanced options so people can make informed choices (e.g. replace ‘you should.’ or ‘this will reduce your risk’ with ‘try to ..’ or ‘if you do this, you are more likely to..’).
- Use gender-sensitive language and acknowledge the role of both reproductive partners, where appropriate.
- The phrases ‘health and wellbeing during the childbearing years’, ‘health and wellbeing before pregnancy and parenthood’ and ‘planning for parenthood’ resonated with most public contributors.

##### Public health messaging recommendations

- Use general messaging about the concept of preconception health, combined with more specific messaging on individual health and wellbeing factors.
- Be specific, and ultimately include the ‘what’, ‘who’, ‘why’ and ‘when’ of the message.
- Highlight the immediate benefits for the person, as well as benefits for a potential future pregnancy and child, where relevant.
- Use health and behaviour messages that are realistic (e.g. provide options), and that don’t provoke blame, guilt and stigma.

Topics discussed in more detail included the need for appropriate and relevant educational messages about preconception health, clarity on why it is important beyond being healthy in general, and the use of gender-sensitive language.

### Educational messages

Based on the limited knowledge and understanding of preconception health in the community, public contributors thought communication about preconception health would need to make clear when, why and to whom it is relevant, and what it involves. This could mean that, in addition to general messages relevant to everyone who may become pregnant or a parent, targeted messages may be needed, for example, for men/male partners (including cisgender and transgender men), people with varying pregnancy intentions in the near and more distant future, and people with previous pregnancy loss or with chronic physical and mental health conditions.

### Preconception health-specific messages

When explaining the ‘why’ of a preconception health message, public contributors felt it important to communicate the immediate benefits for the person, as well as benefits for a future pregnancy and child. Preconception health messages could for example build on existing well-known public health messages about healthy eating and stopping smoking, and highlight the additional benefits for pregnancy and a future family. Public contributors also thought, however, that “everyone knows they have to be healthy” and “we don’t need another reason to be healthy, especially if we’re not thinking of having a baby in the near future”. There are many barriers to healthy eating and stopping smoking (e.g. cost of living crisis), and more health messages would be tiring and frustrating rather than motivating and empowering. On the other hand, public contributors felt that targeted messages might be helpful for people across the childbearing years and at different stages of pregnancy planning, including messages for people who are most likely to have unplanned pregnancies.

### Gender-sensitive language

Given the relevance of preconception health to people of all gender identities, public contributors felt it would be important to reach everyone when recruiting study participants and developing educational and health promotion messages. Not everyone identifies as woman or man, or relates to terms like mother and father. People also agreed most men and younger people would not think that messages that included the word ‘pregnant’ were relevant to them, and rather than ‘planning for pregnancy’ they would relate more to ‘planning for parenthood’. On the other hand, public contributors believed messages that only included gender-neutral terms, such as ‘people who may become pregnant’ or ‘future parents’ would not be specific enough to attract attention of the intended target group. Specification of to whom it is relevant would be needed, for example, by using ‘women and people who may become pregnant’ or ‘women, men and people of other genders who are thinking of having a baby’.

### Case study: “Thinking of having a baby?” poster

The discussions and agreed recommendations on appropriate and relevant language for preconception health were used in Session 2 to improve the language used in an existing poster titled ‘Thinking of having a baby?’.^27^ Changes were made mainly related to recommendations to replace strong and definitive language with more nuanced and realistic options, clarify that all messages are relevant before pregnancy, remove unclear terms, and use gender-sensitive language (**Table 2**). Additional suggestions were made, but due to the limited space on the one-page poster these have not yet been addressed. For example, public contributors were interested in 1) a reference to nutritional supplement recommendations beyond folic acid, 2) further information and signposting to guide, for example, individual food and beverage (e.g. caffeine) choices and smoking cessation support, and 3) more details and explanation on why things are important for sperm health. People liked the use of colour and the way that bold text and underlining draws attention to important details. The numbering of each section was generally well liked, although some contributors were unsure if it implied an order of priority like in a to-do list. It was also considered important that there be no implication that a future pregnancy would definitely be healthy and without complications if all steps included were followed.

**Table 2.** Examples of language changes made to the “Thinking of having a baby?” poster.

| Original text | Reflections | Updated text |
| --- | --- | --- |
| Thinking of having a baby? | The title may only attract those who consider a pregnancy within a relatively short time period.<br>Wider engagement could be encouraged by altering the title to expand the timeframe.<br>“I would not have read past the title for the first 34 years of my life”. | Thinking of having a baby – now or in the near future? |
| It is important for women and for men to be healthy before getting pregnant. | Where possible, gender-sensitive language should be used.<br>Getting pregnant is beyond people’s control. | It is important for women and people able to carry a pregnancy, and their partners, to be as healthy as possible before trying to get pregnant |
| Being healthy before conceiving means a better chance of getting pregnant and a healthier pregnancy. | The term ‘conceiving’ was not understood by everyone.<br>Reference to the health of the baby is missing. | Being healthy before trying to get pregnant means a better chance of getting pregnant and a healthier pregnancy and baby. |
| Taking folic acid for two months before you get pregnant and for the first 3 months of pregnancy lowers the risk of the baby having spina bifida by 70%. | The phrase ‘two months before you get pregnant’ was considered unhelpful as this assumes the timing of (the future) pregnancy is known. | Start taking folic acid about two months before you start trying to get pregnant. Continue to take it for the first 3 months of pregnancy. This lowers the risk of the baby having spina bifida by 70%. |
| Eat more fruit and vegetables (5 portions a day) | Eating 5 portions of fruit and vegetables every day is not realistic for most people. | Eat plenty of fruit and vegetables (aim for 5 portions a day) |
| Quit or reduce smoking | The word ‘quit’ was disliked by smokers in the group, as this was considered ‘impossible’ – suggestion was to start with ‘reduce’ and then mention ‘stop’ | Reduce or stop smoking. |
| Quit alcohol, caffeine and recreational drugs | Inclusion of alcohol, caffeine and recreational drugs in one sentence was considered problematic.<br>“Surely a cup of coffee isn’t as bad as using drugs”. | Quit alcohol & recreational drugs and limit caffeine. |
| Alcohol and recreational drugs can harm the baby and caffeine can increase the risk of miscarriage. | It was unclear if this was relevant before pregnancy. | Alcohol and recreational drugs can harm the baby and too much caffeine before pregnancy can increase the risk of |
|  |  | miscarriage. |
| Speak to your GP several months before getting pregnant if...<br>You have any existing health conditions or are taking regular medications. | • It was unclear if this was relevant only to people who can carry a pregnancy or also to their partner.<br>• Reference to both mental and physical health was considered very important. | Speak to your GP several months before you or your partner are trying to get pregnant if...<br>You have any existing physical or mental health conditions or are taking regular medications. |
| Good for sperm health. | • References to sperm health were very well received, but it was felt that the placement at the end of each relevant section, immediately after negative references, could suggest that these negative things were actually good for sperm health. | Important for sperm health. |

### Approaches to raising understanding and awareness

In Session 1, a range of platforms and settings were identified by public contributors that could be used to increase understanding and raise awareness of preconception health:

- Education in schools, from initial Relationships and Sex Education in secondary schools through to universities, with repeated and updated relevant messages
- Social media (campaigns, including short interactive elements such as videos that feature personal stories)
- Magazines and news articles
- Packaging, for example for period products or contraception (pills and condoms), or smoking and alcohol packaging
- Posters in public places, such as toilets in cinemas, bus and train stations, universities
- Digital screens, posters and brochures at healthcare services and practices
- Information from and conversations with healthcare professionals (GP, nurse, sexual and reproductive health doctor, pharmacist, midwife)
- Information as part of for example exercise or weight loss apps
- Peer-support and community groups with parents and parents-to-be
- Learning from own parents and other family relatives and friends
- Workplace support, for example through ‘wellness hubs’.

In Session 2, thoughts on these ideas that came up in the first session were further discussed and prioritised.

Public contributors felt it would be important to prioritise approaches that would reach as many (diverse) groups of people as possible without the need ‘to proactively do, search or pay for something’, given the general lack of knowledge, understanding and awareness of preconception health. This approach would help normalise conversations about planning and preparing for pregnancy and parenthood, and encourage community support, for example, when people experience pregnancy loss, mental health issues, or other difficulties. These wide-reaching approaches would include education in schools and social media. There was also agreement that preconception health messages should come from and be endorsed by the National Health Service (NHS) as the most trusted source of health information. For example, social media posts and posters that include an NHS logo would be trusted and taken seriously amongst the many health messages promoted through social media. An online NHS information hub that brings together all relevant information in one place was recommended and could be signposted using a wide range of messages distributed in schools, through social media, healthcare settings and other platforms. In addition to online resources, healthcare professionals also have an important role in raising awareness by creating a safe and inclusive space to have respectful conversations about pregnancy and family planning.

Based on these discussions, public contributors agreed the most important strategy would be preconception health education and support through three key routes: 1) schools; 2) social media/public health campaigns; and 3) the NHS.

### Evaluation and feedback

Findings from an anonymous poll at the end of each discussion meeting in Session 1 showed that public contributors thought the meeting invitation and instructions were helpful and adequate and that the facilitator knew their subject well and helped to discuss ideas (Supplementary File 1). All participants strongly agreed that they were glad they participated in the activity. Public contributors were very engaged and keen to learn and share their thoughts and ideas. Sometimes they would have liked more time for discussion; this was also reflected in feedback provided through an anonymous Feedback Form (Supplementary File 1).

## Discussion

This public consultation involving a diverse group of people of reproductive age in the UK confirmed a general lack of knowledge of preconception health, and identified a desire to learn more and contribute to research and development of approaches that raise awareness. Through a series of online discussions, language and public health messaging recommendations were developed that can be used to communicate about preconception health in ways that are informative, relevant and appropriate to the public. Approaches to improving knowledge and understanding of preconception health were identified and prioritised, suggesting school-based education, social media campaigns and NHS resources need to be co-developed to raise public awareness.

A general lack of knowledge and awareness of preconception health among the public has been reported in previous studies.^18–23^ When people are not prompted about what might be important before pregnancy, topics most mentioned include: diet and (folic acid/vitamin) supplements, weight, alcohol, smoking, illicit drugs/substances, medical conditions, wellbeing/mental health, social support and relationships, and financial circumstances.^18, 20, 23^ These topics were not necessarily mentioned or understood as important by the majority of participants in our consultation or previous studies, and detailed knowledge of why these topics are important and what to do (e.g. in relation to dosage and timing of folic acid supplement use) was limited.^18, 22^ Importantly, our consultation confirmed previous findings that people want to learn more,^22^ and consider preconception health to be important, once they know what it is.^28, 29^ This highlights the need to further involve the public in the co-development of interventions that increase knowledge and awareness of preconception health.

Building on what the public know and do not know about preconception health, it is important to consider what they want to know and how this can be best communicated. Discussions throughout our public consultation indicated that people want detailed information on all aspects of preconception health related to whom should be advised to do what, why and when. This may reflect their general lack of knowledge of preconception health,^18–23^ and their willingness to learn more.^22^ Public contributors in our consultation also agreed that they wanted to know about immediate benefits of optimising preconception health for their own health, in addition to benefits for a healthy pregnancy and baby. They noted that most people of reproductive age would not be actively planning pregnancy at a given time or may be unsure about their future parenthood aspirations. This suggests language and educational messages about the relevance and importance of preconception health need to be inclusive of the multiple phases that people move through during their reproductive years in relation to their goal to become, or not become, a parent.^30–32^

Information on all aspects of preconception health should be communicated through research study participant materials and public health messages, to ensure it resonates with people of reproductive age. Public contributors in our consultation concluded that general messages and recruitment text that are relevant to ‘everyone’ of reproductive age would be important to raise widespread awareness and normalise the concept of preparing for pregnancy and parenthood in the community. In addition, targeted messages for specific groups would be needed to attract attention and educate groups of people who may otherwise believe the more general messages are not relevant to them (e.g. men, young people) or who may require tailored information and advice (e.g. specific to cultural beliefs, long-term health conditions). People’s intention to start or grow a family in the near future is likely the strongest influence on their preconception health information receptiveness and needs,^25, 30, 32, 33^ and further research is needed to co-develop public health messages that motivate health and support-seeking behaviour change among diverse groups of people across their reproductive years.

The language that is used in research studies and public health messages is also important to ensure information is communicated in ways that are accessible and appropriate. In line with our findings, previous research has found that the term ‘preconception health’ is not understood by most people of reproductive age;^32, 34^ however, alternative language has not been explored before. Public contributors in our consultation agreed that an alternative short phrase that captures the full concept of preconception health does not exist and may not be needed if people are educated about the term so its meaning becomes clear, normalised and accepted. Education that uses the term ‘preconception health’ may therefore be needed, in addition to the use of alternative phrases that may be more attractive, appropriate and understandable for a broad audience. These include ‘health and wellbeing before pregnancy and parenthood’ and ‘planning for parenthood’.

In addition to relevant and easy-to-understand terminology, the recommendations developed during our public consultation suggest language should be positively framed (focused on benefits rather than risks) and non-judgmental (avoiding blame, guilt and stigma). Many aspects of preconception health are linked to stigma and not easily modifiable (e.g. weight, alcohol consumption, older age). Public health messages about these topics need to be carefully worded and consider potential barriers to behaviours change. These recommendations are in line with a previous qualitative study exploring women’s views on preconception health intervention content,^24^ and learnings from an international collaboration on fertility education and awareness.^35^

The use of gender-sensitive language was also discussed extensively in our public consultation. In line with previous opinion pieces and reviews,^36–38^ our discussions suggest that information and public health messages about preconception health should not use de-sexed language to avoid confusion and inaccuracy related to whom the messages are relevant to, and to address sex– and gender-specific preconception health risk factors and needs. A gender-additive approach was preferred among public contributors where gender-neutral language is used alongside sex-or gender-specific language to ensure that everyone is represented and included (e.g. ‘women and people who may become pregnant’ or ‘women, men and people of other genders who are thinking of having a baby’).

A large variety of platforms and settings were identified in our public consultation as means of general education and awareness of preconception health. In line with other studies,^23, 25, 39^ public contributors in our consultation prioritised education in schools, social media/public health campaigns and healthcare settings/professionals. The consistency and trustworthiness of information was considered important, and public health messages should therefore be consistent, reinforced and endorsed by a well-known (healthcare) organisation (such as the NHS) across multiple relevant platforms and settings.^24, 25^ These priority approaches confirm our previously developed evidence-based framework for integrated contraception and preconception care, proposing awareness raising through education in schools and colleges, social media campaigns and training of health professionals, alongside opportunistic signposting in primary care.^40^ While individual approaches have been developed and tested at local levels,^14, 41–43^ further co-development of a comprehensive preconception health education ‘package’ – with language that is relevant and acceptable in different settings – is now needed to increase public knowledge and understanding of preconception health.

Findings from our public consultation provide new insights from people of reproductive age in the UK into what they want to know and how this can be best communicated to improve public understanding and awareness of preconception health. Our findings should be interpreted in the context of potential limitations. The majority of public contributors were recruited through Southampton-based PPI and community groups and a local charity. Views were therefore mostly from public contributors living in the Southampton area, even though the group included people with diverse characteristics and from all UK nations. People of reproductive age younger than 18 years (i.e. those aged 15-17 years) were not included and may have different opinions on relevant language use and preferred approaches to raise awareness. This group may be particularly relevant to include in future consultations if education on preconception health is to take place in schools. No formal data were collected on, for example, people’s socioeconomic status, ethnicity, health literacy and previous pregnancy experiences and future pregnancy plans, so we were not able to explore if people’s views differed based on these characteristics. We only explored communication about preconception health in the English language, and further public consultations are needed to develop recommendations appropriate for other languages.

## Conclusion

People of reproductive age in the UK have little understanding of preconception health. More needs to be done to effectively engage the public in research and interventions that support planning and preparation for pregnancy and parenthood. The language recommendations developed in this public consultation can be used in future preconception health research, including study participant materials and dissemination of research findings, and to inform further co-development of school-based education, social media campaigns and NHS-based interventions to raise awareness in ways that are relevant and appropriate to the public.

## Data availability

The data that supports the findings of this public consultation are available in the article or in its supplementary material.

## Ethics approval

Ethics approval was not required for this public consultation.

## Acknowledgements

The authors thank all public contributors for sharing their views during the public consultation activities, and for their ongoing involvement in our research.

## Supplementary File 1

### 1) Anonymous evolution poll results from session 1, N = 54 public contributors

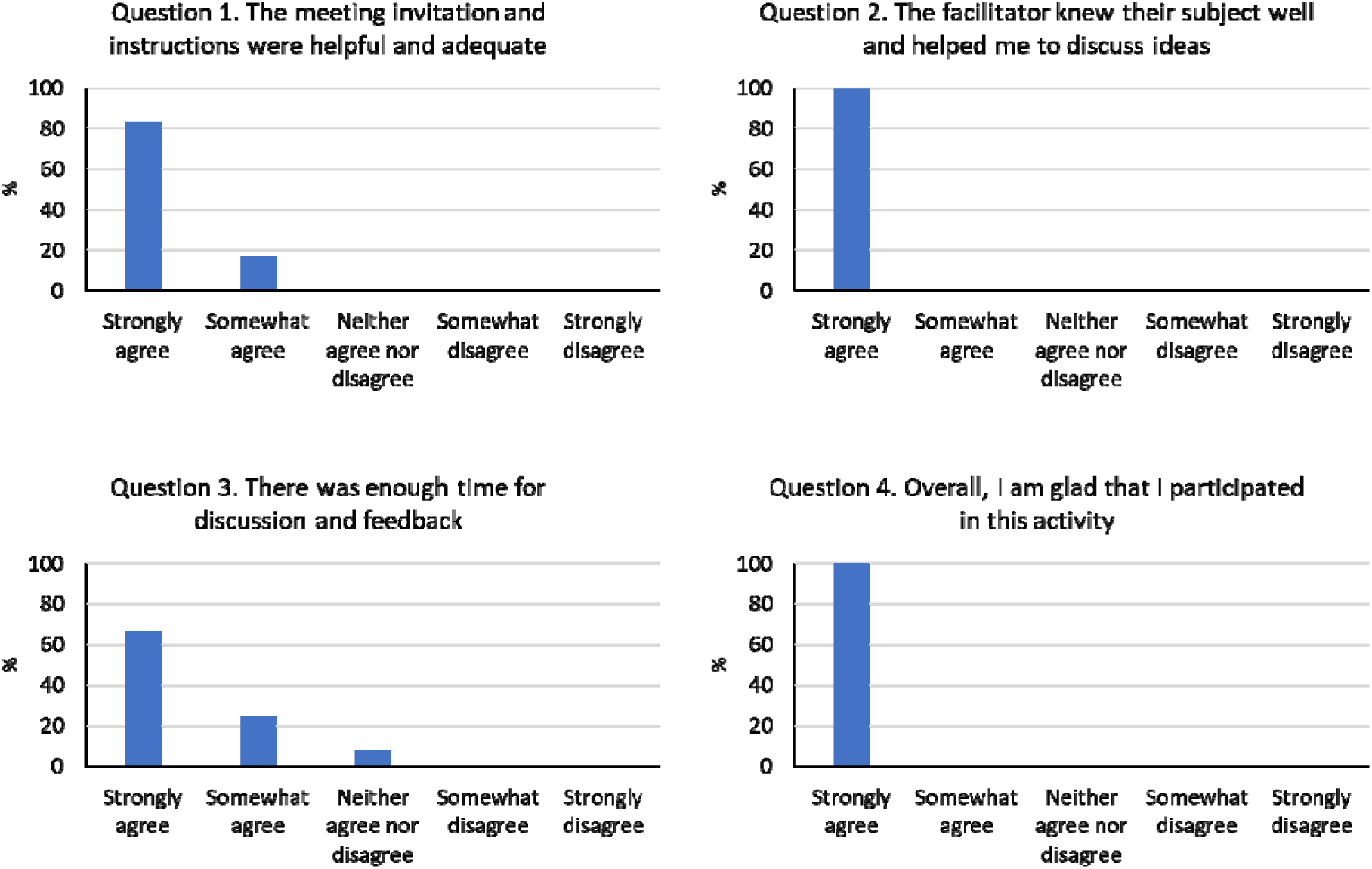

### 2) Anonymous Feedback Form results from session 1, N = 32 public contributors

Selected quotes to cover common themes:

<u>Please provide any feedback about what you enjoyed or found useful about this activity</u>

“There was no pressure and the conversation flowed well. Everyone got a chance to say something and the facilitator was very knowledgeable and engaging and made me feel like my contribution was valuable.”

“I was really intrigued on the subject. I hadn’t realised that our health from many years before pregnancy would make an impact on the ability to get pregnancy, and more so the complications during childbirth and pregnancy.”

“Great to meet everyone, there was so much to learn from other people’s experiences and thoughts.”

“Among everyone I don’t think there was a master on the subject, but they all made salient points. I really enjoyed the discussions as they provided me with things I didn’t know before!”

<u>Is there anything you think could be improved?</u>

“A little more time would have been good.”

“Might have been useful to have some examples of the key messages as they stand.”

“Specific examples of wording that is currently being used and the context it’s used in, as well as examples of potential other wording.”

“There was some repetition as some questions were asked multiple times. Although I guess this was to involve everyone in the discussion, those who hadn’t spoken yet.”

<u>Is there anything that you will take away as a result of the discussion?</u>

“Made me think about my own preconception health, and the effects my choices will have on my body later in life.”

“I now have a different perspective to the importance of health before having children and the discussion has made me recognise the lack of education in this area.”

“I learned that the health of the male before pregnancy can impact the health of the baby too! More information on this is needed.”

“The discussion on positive and motivational approaches to promotion of health before pregnancy was really interesting. Also, the importance of language used in messages and how these may affect information uptake.”

<u>Please use the space below to comment on any other part of the activity</u>

“Thank you so much for the opportunity to be involved in such an interesting discussion! I plan to spend the voucher on some books on health to keep learning, so thank you so much!”

“Really interesting! Thanks! Looking forward to being further involved.”

## References

1. World Health Organization (WHO). Preconception care: Maximizing the gains for maternal and child health – Policy brief. Geneva, World Health Organization, 2013. Available from: https://www.who.int/publications/i/item/WHO-FWC-MCA-13.02. [accessed 04/06/2024].

2. Stephenson J, Heslehurst N, Hall J, Schoenaker D, Hutchinson J, Cade JE, et al. Before the beginning: nutrition and lifestyle in the preconception period and its importance for future health. Lancet. 2018;391(10132):1830–41.

3. De-Regil LM, Peña-Rosas JP, Fernández-Gaxiola AC, Rayco-Solon P. Effects and safety of periconceptional oral folate supplementation for preventing birth defects. Cochrane Database Syst Rev. 2015;2015(12):Cd007950.

4. Caut C, Schoenaker D, McIntyre E, Vilcins D, Gavine A, Steel A. Relationships between Women’s and Men’s Modifiable Preconception Risks and Health Behaviors and Maternal and Offspring Health Outcomes: An Umbrella Review. Semin Reprod Med. 2022;40(3-04):170–83.

5. Carter T, Schoenaker D, Adams J, Steel A. Paternal preconception modifiable risk factors for adverse pregnancy and offspring outcomes: a review of contemporary evidence from observational studies. BMC Public Health. 2023;23(1):509.

6. Righton O, Flynn A, Alwan N, Schoenaker D. Preconception health in adolescence and adulthood across generations in the UK: findings from three British birth cohort studies. medRxiv [preprint]. 2024. doi: 10.1101/2024.02.06.24302400.

7. McDougall B, Kavanagh K, Stephenson J, Poston L, Flynn AC, White SL. Health behaviours in 131,182 UK women planning pregnancy. BMC Pregnancy Childbirth. 2021;21(1):530.

8. Stewart C, Hall J. Pregnancy preparation among women and their partners in the UK: how common is it and what do people do? Women’s Reproductive Health. 2023. Doi: 10.1080/23293691.2023.2271919.

9. Schoenaker D, Stephenson J, Smith H, Thurland K, Duncan H, Godfrey KM, et al. Women’s preconception health in England: a report card based on cross-sectional analysis of national maternity services data from 2018/2019. BJOG. 2023;130(10):1187–95.

10. Wellings K, Jones KG, Mercer CH, Tanton C, Clifton S, Datta J, et al. The prevalence of unplanned pregnancy and associated factors in Britain: findings from the third National Survey of Sexual Attitudes and Lifestyles (Natsal-3). Lancet. 2013;382(9907):1807-16.

11. Hall JA, Stewart C, Stoneman B, Bicknell T, Lovell H, Duncan H, et al. Implementation of the London Measure of Unplanned Pregnancy in routine antenatal care: A mixed-methods evaluation in three London NHS Trusts. Eur J Midwifery. 2024;8.

12. Cassinelli EH, McKinley MC, Kent L, Eastwood KA, Schoenaker D, Trew D, et al. Preconception health and care policies, strategies and guidelines in the UK and Ireland: a scoping review. BMC Public Health. 2024;24(1):1662.

13. Dorney E, Boyle JA, Walker R, Hammarberg K, Musgrave L, Schoenaker D, et al. A Systematic Review of Clinical Guidelines for Preconception Care. Semin Reprod Med. 2022;40(3-04):157–69.

14. NHS South East Clinical Delivery and Networks. Ready for Pregnancy campaign. Available from: https://www.southeastclinicalnetworks.nhs.uk/readyforpregnancy/. [accessed 04/06/2024].

15. Bedfordshire, Luton and Milton Keynes Integrated Care System. Planning for Pregnancy. Available from: https://www.healthwatchbedfordborough.co.uk/planning-pregnancy. [accessed 04/06/2024].

16. NHS Lanarkshire. Pre-Pregnancy Health. Available from: https://www.lanarkshiresexualhealth.org/pre-pregnancy-health/. [accessed 04/06/2024].

17. Stevens A, Connolly A. Delivering innovative health interventions to inclusion health populations: lessons learnt from a preconception care programme. Available from: https://bjgplife.com/delivering-innovative-health-interventions-to-inclusion-health-populations-lessons-learnt-from-a-preconception-care-programme/. [accessed 04/06/2024].

18. Welshman H, Dombrowski S, Grant A, Swanson V, Goudreau A, Currie S. Preconception knowledge, beliefs and behaviours among people of reproductive age: A systematic review of qualitative studies. Prev Med. 2023;175:107707.

19. Cairncross ZF, Ravindran S, Yoganathan S, Dennis CL, Enders J, Graves L, et al. Measurement of Preconception Health Knowledge: A Systematic Review. Am J Health Promot. 2019;33(6):941–54.

20. Rabiei Z, Shariati M, Mogharabian N, Tahmasebi R, Ghiasi A, Motaghi Z. Men’s knowledge of preconception health: A systematic review. J Family Med Prim Care. 2023;12(2):201–7.

21. Schoenaker D, Hall J, Stewart S, Hanley SJ, Cassinelli EH, Benton M, Wynn-Jones AA, Chawla M, Currie S, 2023 UK Preconception EMCR Network conference workshop participants. Tackling inequalities in preconception health and care: barriers, facilitators and recommendations for action from the 2023 UK Preconception EMCR Network conference. medRxiv [preprint]. 2024. doi: 10.1101/2024.02.13.24302690v1.

22. McGowan L, Lennon-Caughey E, Chun C, McKinley MC, Woodside JV. Exploring preconception health beliefs amongst adults of childbearing age in the UK: a qualitative analysis. BMC Pregnancy Childbirth. 2020;20(1):41.

23. Daly MP, White J, Sanders J, Kipping RR. Women’s knowledge, attitudes and views of preconception health and intervention delivery methods: a cross-sectional survey. BMC Pregnancy Childbirth. 2022;22(1):729.

24. Daly MP, Kipping RR, White J, Sanders J. Women’s views on content and delivery methods for interventions to improve preconception health: a qualitative exploration. Front Public Health. 2024;12:1303953.

25. Walker R, Drakeley S, Boyle J. Preconception women’s views of promoting preconception women’s health in Australia. Health Promot J Austr. 2021;32 Suppl 2:22–8.

26. Goodfellow A, Frank J, McAteer J, Rankin J. Improving preconception health and care: a situation analysis. BMC Health Serv Res. 2017;17(1):595.

27. Contraception Choices (website). Available from https://www.contraceptionchoices.org/did-you-know/thinking-having-baby. [accessed 04/06/2024].

28. Daly M, Kipping RR, Tinner LE, Sanders J, White JW. Preconception exposures and adverse pregnancy, birth and postpartum outcomes: Umbrella review of systematic reviews. Paediatr Perinat Epidemiol. 2022;36(2):288–99.

29. Cassinelli EH, McClure A, Cairns B, Griffin S, Walton J, McKinley MC, et al. Exploring Health Behaviours, Attitudes and Beliefs of Women and Men during the Preconception and Interconception Periods: A Cross-Sectional Study of Adults on the Island of Ireland. Nutrients. 2023;15(17).

30. Barker M, Dombrowski SU, Colbourn T, Fall CHD, Kriznik NM, Lawrence WT, et al. Intervention strategies to improve nutrition and health behaviours before conception. Lancet. 2018;391(10132):1853–64.

31. Grace B, Shawe J, Johnson S, Usman NO, Stephenson J. The ABC of reproductive intentions: a mixed-methods study exploring the spectrum of attitudes towards family building. Hum Reprod. 2022;37(5):988–96.

32. Lynch M, Squiers L, Lewis MA, Moultrie R, Kish-Doto J, Boudewyns V, et al. Understanding Women’s Preconception Health Goals: Audience Segmentation Strategies for a Preconception Health Campaign. Soc Mar Q. 2014;20(3):148–64.

33. Walker R, Quong S, Olivier P, Wu L, Xie J, Boyle J. Empowerment for behaviour change through social connections: a qualitative exploration of women’s preferences in preconception health promotion in the state of Victoria, Australia. BMC Public Health. 2022;22(1):1642.

34. Dorney E, K IB, Haas M, Street D, Church J. The preferences of people in Australia to respond and engage with advertisements to promote reproductive health: Results of a discrete choice experiment. Prev Med Rep. 2024;40:102657.

35. Mertes H, Harper J, Boivin J, Ekstrand Ragnar M, Grace B, Moura-Ramos M, et al. Stimulating fertility awareness: the importance of getting the language right. Hum Reprod Open. 2023;2023(2):hoad009.

36. Gribble KD, Bewley S, Bartick MC, Mathisen R, Walker S, Gamble J, et al. Effective Communication About Pregnancy, Birth, Lactation, Breastfeeding and Newborn Care: The Importance of Sexed Language. Front Glob Womens Health. 2022;3:818856.

37. Knight M, Nelson-Piercy C. Clarity of guidelines concerning the care of pregnant women is lost by the use of de-sexed language. Br J Haematol. 2023;202(2):437.

38. Garad RM, Bahri-Khomami M, Busby M, Burgert TS, Boivin J, Teede HJ. Breaking Boundaries: Toward Consistent Gender-Sensitive Language in Sexual and Reproductive Health Guidelines. Semin Reprod Med. 2023;41(1-02):5–11.

39. Musgrave L, Homer C, Gordon A. Knowledge, attitudes and behaviours surrounding preconception and pregnancy health: an Australian cross-sectional survey. BMJ Open. 2023;13(1):e065055.

40. Hall J, Chawla M, Watson D, Jacob CM, Schoenaker D, Connolly A, et al. Addressing reproductive health needs across the life course: an integrated, community-based model combining contraception and preconception care. Lancet Public Health. 2023;8(1):e76–e84.

41. Woods-Townsend K, Hardy-Johnson P, Bagust L, Barker M, Davey H, Griffiths J, et al. A cluster-randomised controlled trial of the LifeLab education intervention to improve health literacy in adolescents. PLoS One. 2021;16(5):e0250545.

42. Charafeddine L, El Rafei R, Azizi S, Sinno D, Alamiddine K, Howson CP, et al. Improving awareness of preconception health among adolescents: experience of a school-based intervention in Lebanon. BMC Public Health. 2014;14:774.

43. Poels M, van Stel HF, Franx A, Koster MPH. The effect of a local promotional campaign on preconceptional lifestyle changes and the use of preconception care. Eur J Contracept Reprod Health Care. 2018;23(1):38–44.

